# Association between Psychotropic Medications Functionally Inhibiting Acid Sphingomyelinase and reduced risk of Intubation or Death among Individuals with Mental Disorder and Severe COVID-19: an Observational Study

**DOI:** 10.1101/2021.02.18.21251997

**Authors:** Nicolas Hoertel, Marina Sánchez-Rico, Erich Gulbins, Johannes Kornhuber, Alexander Carpinteiro, Miriam Abellán, Pedro de la Muela, Raphaël Vernet, Nathanaël Beeker, Antoine Neuraz, Aude Delcuze, Jesús M. Alvarado, Pierre Meneton, Frédéric Limosin, On behalf of AP-HP / Universities / INSERM COVID-19 research collaboration and AP-HP COVID CDR Initiative

## Abstract

Prior preclinical and clinical evidence suggests that the acid sphingomyelinase (ASM)/ceramide system may provide a useful framework for better understanding SARS-CoV-2 infection and the repurposing of psychotropic medications with functional inhibition of acid sphingomyelinase, called FIASMA psychotropic medications, against COVID-19. We examined the potential usefulness of FIASMA psychotropic medication use among patients with mental disorder hospitalized for severe COVID-19, in an observational multicenter retrospective study conducted at AP-HP Greater Paris University hospitals. Of 545 adult patients with mental disorder hospitalized for severe COVID-19, 164 (30.1%) received a psychotropic FIASMA medication at study baseline, which was defined as the date of hospital admission for COVID-19. The primary endpoint was a composite of intubation or death. We compared this endpoint between patients who received a psychotropic FIASMA medication at baseline and those who did not in time-to-event analyses adjusted for sociodemographic characteristics, psychiatric and other medical comorbidity, and psychotropic and other medications. The primary analysis was a Cox regression model with inverse probability weighting (IPW). There was a significant association between FIASMA psychotropic medication use at baseline and reduced risk of intubation or death both in the crude analysis (HR=0.42; 95%CI=0.31-0.57; p<0.01) and in the primary IPW analysis (HR=0.50; 95%CI=0.37-0.67; p<0.01). This association remained significant in multiple sensitivity analyses. Exploratory analyses suggested that this association was not specific to one FIASMA psychotropic class or medication. These results suggest the usefulness of the ASM/ceramide system framework in COVID-19. Double-blind controlled randomized clinical trials of these medications for COVID-19 are needed.

## 1. Introduction

Global spread of the novel coronavirus SARS-CoV-2, the causative agent of coronavirus disease 2019 (COVID-19), has created an unprecedented infectious disease crisis worldwide [1–3]. Although the availability of vaccines has raised hope for a decline of the pandemic, the search for an effective treatment for patients with COVID-19 among all available medications is still urgently needed.

Prior preclinical evidence supports that SARS-CoV-2 activates the acid sphingomyelinase (ASM)/ceramide system, resulting in the formation of ceramide-enriched membrane domains that serve viral entry and infection by clustering ACE2, the cellular receptor of SARS-CoV-2 [4]. An *in vitro* study [4] showed that several FIASMA (Functional Inhibitors of Acid Sphingomyelinase Activity [5]) antidepressant medications, including amitriptyline, imipramine, desipramine, fluoxetine, sertraline, escitalopram and maprotiline, inhibited ASM and the formation of ceramide-enriched membrane domains, and prevented Vero cells from being infected with SARS-CoV-2. Reconstitution of ceramide in cells treated with these FIASMA antidepressant medications restored infection with SARS-CoV-2. Oral use of the FIASMA antidepressant amitriptyline in healthy volunteers also efficiently blocked infection of freshly isolated nasal epithelial cells with SARS-CoV-2 [4]. These preclinical data were confirmed by another study that demonstrated an inhibition of the infection of cultured epithelial cells with SARS-CoV-2 by the FIASMA antidepressant fluoxetine [6].

Findings from two recent clinical studies are consistent with these preclinical results. First, a randomized double-blind controlled study [7] showed significant protective effects of the FIASMA antidepressant fluvoxamine (N=80) versus placebo (N=72) on COVID-19 disease progression in outpatients (absolute difference, 8.7% from the survival analysis; log rank p=0.009). Second, an observational multicenter retrospective study using data from the Assistance Publique-Hôpitaux de Paris (AP-HP) Health Data Warehouse showed that use of antidepressants, mostly FIASMA antidepressants, and the FIASMA hydroxyzine, were significantly associated with reduced mortality in patients hospitalized for COVID-19 [8, 9]. Taken together, these results suggest that the ASM/ceramide system may provide a useful framework for better understanding SARS-CoV-2 infection and the repurposing of FIASMA psychotropic medications, whose short-term use is generally well-tolerated [10, 11], for preventing or treating COVID-19.

Several psychotropic medications, including certain antidepressants (i.e., amitriptyline, clomipramine, desipramine, doxepin, duloxetine, escitalopram, fluoxetine, fluvoxamine, imipramine, lofepramine, maprotiline, nortriptyline, paroxetine, protriptyline, sertraline, and trimipramine), certain antipsychotics (i.e., aripiprazole, chlorpromazine, chlorprothixene, fluphenazine, flupenthixol, penfluridol, perphenazine, pimozide, promazine, sertindole, thioridazin, trifluoperazine, and triflupromazine), and hydroxyzine, have shown to *in vitro* inhibit acid sphingomyelinase with varying degrees [4, 5, 12, 13]. To our knowledge, no clinical study to date has examined the potential usefulness of FIASMA psychotropic medications in patients hospitalized for COVID-19. Observational studies of patients with COVID-19 taking medications for other indications can help decide which treatment should be prioritized for randomized clinical trials and reduce the risk for patients of being exposed to potentially harmful and ineffective treatments [8, 9, 14–16].

In this report, we used data from the AP-HP Health Data Warehouse and examined the association between FIASMA psychotropic medication use and the composite outcome of intubation or death among patients with mental disorder hospitalized for severe COVID-19. We focused on this population because individuals with mental disorder are a vulnerable population at higher risk of severe COVID-19 [9, 17, 18] likely to receive psychotropic medications for treating or preventing the relapse of psychiatric disorders. If a significant protective association were found, we sought to perform additional exploratory analyses to examine whether this association was specific to certain FIASMA psychotropic classes (e.g., antidepressants) or individual medications (e.g., fluoxetine). Our primary hypothesis was that FIASMA psychotropic medication use would be associated with reduced risk of intubation or death among patients with mental disorder hospitalized for severe COVID-19 in time-to-event analyses adjusting for sociodemographic characteristics, psychiatric and other medical comorbidity, and psychotropic and other medications. If this were the case, our secondary hypothesis was that this association would not be specific to one FIASMA psychotropic class or medication.

## 2. Methods

### 2.1. Setting and Cohort Assembly

We conducted a multicenter observational retrospective cohort study at 36 AP-HP hospitals from the beginning of the epidemic in France, i.e. January 24^th^, until May 1^st^. We included all adults aged 18 years or over with a mental disorder who have been hospitalized in these medical centers for severe COVID-19. Mental disorder was defined as having at least one current International Statistical Classification of Diseases and Related Health Problems (ICD-10) diagnosis of mental disorder (F01-F99) during the visit or an ongoing prescription of any antidepressant, antipsychotic, or mood stabilizer (i.e. lithium or antiepileptic medications with mood stabilizing effects) for a psychiatric disorder at hospital admission. COVID-19 was ascertained by a positive reverse-transcriptase–polymerase-chain-reaction (RT-PCR) test from analysis of nasopharyngeal or oropharyngeal swab specimens. Severe COVID-19 was defined as having at least one of the following criteria at baseline [19–21]: respiratory rate >24 breaths/min or <12 breaths/min, resting peripheral capillary oxygen saturation in ambient air <90%, temperature >40°C, systolic blood pressure <100 mm Hg, lactate levels >2 mmol/L, or admission to an intensive care unit (ICU) within the first 24 hours form hospital admission.

This observational study using routinely collected data received approval from the Institutional Review Board of the AP-HP clinical data warehouse (decision CSE-20-20_COVID19, IRB00011591). AP-HP clinical Data Warehouse initiatives ensure patient information and informed consent regarding the different approved studies through a transparency portal in accordance with European Regulation on data protection and authorization n°1980120 from National Commission for Information Technology and Civil Liberties (CNIL). All procedures related to this work adhered to the ethical standards of the relevant national and institutional committees on human experimentation and with the Helsinki Declaration of 1975, as revised in 2008.

### 2.2. Data sources

We used data from the AP-HP Health Data Warehouse (‘Entrepôt de Données de Santé (EDS)’). This warehouse contains all available clinical data on all inpatient visits for COVID-19 to 36 Greater Paris University hospitals. The data included patient demographic characteristics, vital signs, laboratory test and RT-PCR test results, medication administration data, medication lists during current and past hospitalizations in AP-HP hospitals, current diagnoses, discharge disposition, and death certificates.

### 2.3. Variables assessed

We obtained the following data for each patient at the time of the hospitalization: sex; age; hospital; obesity; current smoking status; any medication prescribed according to compassionate use or as part of a clinical trial; current psychiatric disorder (i.e. ICD-10 diagnosis of substance use disorder, psychotic disorder, mood or anxiety disorder, delirium or dementia, and other psychiatric disorders); and any prescription for antidepressant, mood stabilizer, benzodiazepine or Z-drug, or antipsychotic medication. These variables are detailed in **Supplementary Text**.

### 2.4. Psychotropic medications functionally inhibiting acid sphingomyelinase (ASM)

FIASMA psychotropic medications were defined as psychotropic medications showing a substantial *in vitro* functional inhibition effect on ASM (i.e., a residual ASM activity lower than 50%), as described in detail elsewhere [4, 5, 12, 13]. FIASMA psychotropic medication use was defined as receiving at least one psychotropic FIASMA medication at study baseline, i.e. within the first 24 hours of hospital admission, and before the end of the index hospitalization, intubation or death. To minimize potential confounding effects of late prescription of FIASMA psychotropic medications, patients who received a FIASMA psychotropic medication 24 hours after hospital admission were excluded from the analyses. Finally, patients who received at study baseline any antipsychotic in ICU, possibly as an aid to oral intubation, were also excluded.

### 2.5. Primary endpoint

Study baseline was defined as the date of hospital admission for COVID-19. The primary endpoint was the time from study baseline to intubation or death. For patients who died after intubation, the timing of the primary endpoint was defined as the time of intubation. Patients without an end-point event had their data censored on May 1^st^, 2020.

### 2.6. Statistical analysis

We calculated frequencies of baseline characteristics described above in patients receiving or not receiving a FIASMA psychotropic medication and compared them using standardized mean differences (SMD).

To examine the association between FIASMA psychotropic medication use and the endpoint of intubation or death, we performed Cox proportional-hazards regression models [22]. To help account for the nonrandomized prescription of psychotropic medications and reduce the effects of confounding, the primary analysis used propensity score analysis with inverse probability weighting (IPW) [23, 24]. Given the expected relatively limited sample size and the number of potentially influencing variables, a backward stepwise Cox regression was used to assess the importance of the covariates (listed in **Supplementary Table 1**) on the outcome, based on clinical meaningfulness and the Akaike Information Criterion (AIC) for model comparison [25]. Next, the individual propensities for receiving a FIASMA psychotropic medication were estimated using a multivariable logistic regression model including the variables from the model with the lowest AIC value. In the inverse-probability-weighted analyses, the predicted probabilities from the propensity-score models were used to calculate the stabilized inverse-probability-weighting weights [23]. The association between FIASMA psychotropic medication use and the endpoint was then estimated using an IPW Cox regression model. In case of unbalanced covariates, an IPW multivariable Cox regression model adjusting for the unbalanced covariates was also performed. Kaplan-Meier curves were performed using the inverse-probability-weighting weights [26, 27] and their pointwise 95% confidence intervals were estimated using the nonparametric bootstrap method [27].

We conducted two sensitivity analyses. First, we performed a multivariable Cox regression model including as covariates the same variables used in the IPW analysis. Second, we used a univariate Cox regression model in a matched analytic sample using a 1:1 ratio, based on the same variables used for the IPW analysis and the multivariable Cox regression analysis. To reduce the effects of confounding, optimal matching was used in order to obtain the smallest average absolute distance across all clinical characteristics between exposed patients and non-exposed matched controls [28].

We performed four additional exploratory analyses. First, we examined the relationships between each FIASMA psychotropic class (i.e. FIASMA antidepressants and antipsychotics) and each individual FIASMA molecule with the endpoint. Second, we examined within each psychotropic class (i.e. antidepressants and antipsychotics) the relationships of FIASMA and non-FIASMA molecules with the endpoint. Third, because of discrepancies in the potential FIASMA *in vitro* effect of venlafaxine, mirtazapine, and citalopram [4, 5], we reproduced the main analyses while considering these molecules as FIASMAs. Finally, we reproduced the main analyses among all patients with mental disorder with and without clinical severity criteria at baseline.

For all associations, we performed residual analyses to assess the fit of the data, check assumptions, including proportional hazards assumption using proportional hazards tests and diagnostics based on weighted residuals [22, 29], and examined the potential influence of outliers. Because our main analysis focused on the association between FIASMA psychotropic medication use and the composite outcome of intubation or death among patients with mental disorder hospitalized for severe COVID-19, statistical significance was fixed *a priori* at two-sided p-value <0.05. Only if a significant protective association were found, we planned to perform additional exploratory analyses as described above. All analyses were conducted in R software version 2.4.3 (R Project for Statistical Computing).

## 3. Results

### 3.1. Characteristics of the cohort

Of the 17,131 patients with a positive COVID-19 RT-PCR test who had been hospitalized for COVID-19, 1,963 (11.5%) were excluded because of missing data or their young age (i.e. less than 18 years old of age). Of 15,168 adult inpatients, 1,998 (13.2%) had a mental disorder diagnosis or an ongoing prescription of any antidepressant, antipsychotic, or mood stabilizer at hospital admission. Of these 1,998 patients, 827 (41.4%) had criteria for severe COVID-19. Of these 827 patients, 281 (34.0%) were excluded because they received a FIASMA psychotropic medication after 24 hours from hospital admission (N=277) or because they received an antipsychotic at baseline while being hospitalized in an ICU, possibly as an aid for intubation (N=5). Of the remaining 545 adult inpatients with mental disorder and severe COVID-19, 164 (30.1%) received a FIASMA psychotropic medication at baseline and 381 (69.9%) did not (**Figure 1**).

**Figure 1.**
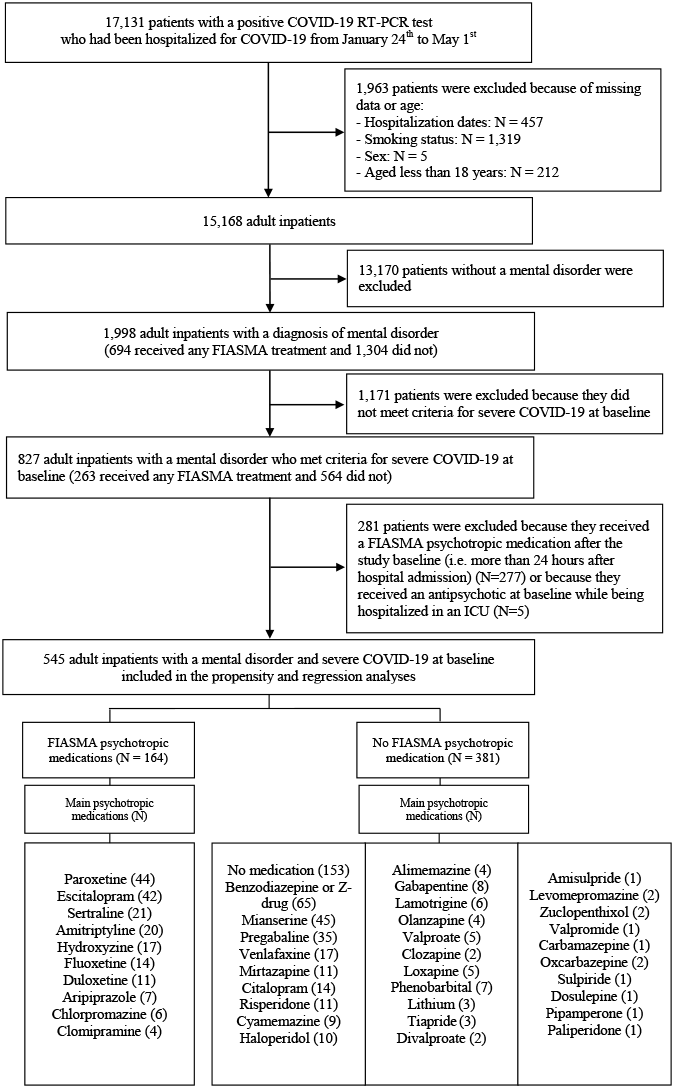
Study cohort.

PT-PCR test results were obtained after a median delay of 0.9 days (SD=9.4) from hospital admission date. This median delay was 0.9 days in the exposed group (SD=11.4) and the non-exposed group (SD=9.3).

Over a mean follow-up of 9.2 days (SD=12.5; median=6 days), 272 patients (50.0%) had an end-point event at the time of data cutoff on May 1^st^. Among patients who received a FIASMA psychotropic medication at baseline, the mean follow-up was 12.0 days (SD=12.9, median=8 days), while it was of 8.9 days (SD=12.4, median=5 days) in those who did not.

Sex, hospital, number of medical conditions, delirium or dementia, any other mental disorder, and the prescription of any antidepressant, any antipsychotic, and any mood stabilizer were significantly associated with the endpoint of intubation or death (**Supplementary Table 1**). A backward stepwise Cox regression showed that a model including age, sex, hospital, obesity, and the number of medical conditions, was meaningful and associated with the lowest AIC value (**Supplementary Table 2**).

The distributions of patient characteristics included in the propensity and regression analyses according to FIASMA psychotropic medication use are shown in **Table 1**. In the full sample, FIASMA psychotropic medication use substantially differed according to age, sex, hospital, and number of medical conditions. After applying the propensity score weights, there were no differences (i.e. all SMD<0.1) in any characteristic. In the matched analytic sample using a 1:1 ratio, sex, and the number of medical conditions differed between groups (**Table 1**).

**Table 1.** Characteristics of patients with mental disorder and severe COVID-19 receiving or not receiving FIASMA psychotropic medications at baseline (N=545).

|  | Exposed to any<br>FIASMA<br>(N=164) | Not exposed to<br>any FIASMA<br>(N=381) | Non-exposed<br>matched group<br>(N=164) | Exposed to any<br>FIASMA<br>vs.<br>Not exposed | Exposed to any FIASMA<br>vs.<br>Not exposed | Exposed to any<br>FIASMA<br>vs.<br>Non-exposed<br>matched group |
| --- | --- | --- | --- | --- | --- | --- |
|  |  |  |  | Crude analysis | Analysis weighted by<br>inverse-probability-weighting weights | Matched analytic<br>sample analysis |
|  | N (%) | N (%) | N (%) | SMD | SMD | SMD |
| Age |  |  |  | **0.197** | 0.037 | 0.022 |
| 18 to 50 years | 19 (11.6%) | 26 (6.82%) | 18 (48.6%) |  |  |  |
| 51 to 70 years | 43 (26.2%) | 105 (27.6%) | 43 (50.0%) |  |  |  |
| 71 to 80 years | 37 (22.6%) | 75 (19.7%) | 38 (50.7%) |  |  |  |
| More than 80 years | 65 (39.6%) | 175 (45.9%) | 65 (50.0%) |  |  |  |
| Sex |  |  |  | **0.260** | 0.008 | **0.159** |
| Women | 90 (54.9%) | 160 (42.0%) | 77 (46.1%) |  |  |  |
| Men | 74 (45.1%) | 221 (58.0%) | 87 (54.0%) |  |  |  |
| Hospital |  |  |  | **0.267** | 0.024 | 0.068 |
| AP-HP Centre – Paris University,<br>Henri Mondor University Hospitals and<br>at home hospitalization | 36 (22.0%) | 115 (30.2%) | 40 (52.6%) |  |  |  |
| AP-HP Nord and Hôpitaux<br>Universitaires Paris Seine-Saint-Denis | 45 (27.4%) | 77 (20.2%) | 46 (50.5%) |  |  |  |
| *AP-HP Paris Saclay University* | 41 (25.0%) | 113 (29.7%) | 38 (48.1%) |  |  |  |
| *AP-HP Sorbonne University* | 42 (25.6%) | 76 (19.9%) | 40 (48.8%) |  |  |  |
| Obesity <sup>1</sup> |  |  |  | **0.129** | 0.001 | **0.162** |
| *Yes* | 42 (25.6%) | 77 (20.2%) | 31 (42.5%) |  |  |  |
| *No* | 122 (74.4%) | 304 (79.8%) | 133 (52.2%) |  |  |  |
| Number of medical conditions <sup>2</sup> |  |  |  | **0.321** | 0.015 | 0.029 |
| *0* | 42 (25.6%) | 51 (13.4%) | 40 (48.8%) |  |  |  |
| *1* | 17 (10.4%) | 37 (9.71%) | 17 (50.0%) |  |  |  |
| *2 or more* | 105 (64.0%) | 293 (76.9%) | 107 (50.5%) |  |  |  |

<sup>1</sup> Defined as having a body-mass index higher than 30 kg/m<sup>2</sup> or an International Statistical Classification of Diseases and Related Health Problems (ICD-10) diagnosis code for obesity (E66.0, E66.1, E66.2, E66.8, E66.9).
<sup>2</sup> Assessed using ICD-10 diagnosis codes for diabetes mellitus (E11), diseases of the circulatory system (I00-I99), diseases of the respiratory system (J00-J99), neoplasms (C00-D49), diseases of the blood and blood-forming organs and certain disorders involving the immune mechanism (D5-D8), frontotemporal dementia (G31.0), peptic ulcer (K27), diseases of liver (K70-K95), hemiplegia or paraplegia (G81-G82), acute kidney failure or chronic kidney disease (N17-N19), and HIV (B20).
SMD>0.1 indicate substantial differences (bold).
Abbreviation: SMD, standardized mean difference.

### 3.2. Study endpoint

The endpoint of death occurred in 57 patients (34.8%) who received a FIASMA psychotropic medication at baseline and 215 patients (56.4%) who did not. The crude, unadjusted analysis (hazard ratio (HR)=0.42; 95% CI=0.31-0.57; p<0.001) and the primary analysis with inverse probability weighting (HR=0.50; 95% CI=0.37-0.67; p<0.001) showed a significant association between FIASMA psychotropic medication use and reduced risk of intubation or death (**Figure 2**; **Table 2**). A post-hoc analysis indicated that we had 80% power in the crude analysis to detect a hazard ratio of at least 0.60 / 1.71.

**Table 2.** Association between FIASMA psychotropic medication use at baseline and risk of intubation or death among patients with mental disorder hospitalized for severe COVID-19.

|  | Number of events<br>/ Number of<br>patients | Crude Cox<br>regression analysis | Multivariable Cox<br>regression<br>analysis<sup>1</sup> | Analysis weighted<br>by inverse-probability-weighting weights<sup>1</sup> | Number of<br>events /<br>Number of<br>patients in the<br>matched groups | Univariate Cox<br>regression in a 1:1<br>ratio matched<br>analytic sample<sup>1</sup> | Cox regression in<br>a 1:1 ratio<br>matched analytic<br>sample<sup>1</sup> adjusted<br>for unbalanced<br>covariates<sup>2</sup> |
| --- | --- | --- | --- | --- | --- | --- | --- |
|  | N (%) | HR (95% CI; p-value) | HR (95% CI; p-value) | HR (95% CI; p-value) | N (%) | HR (95% CI; p-value) | HR (95% CI; p-value) |
| No FIASMA<br>psychotropic<br>medication | 215 / 381 (56.4%) | Ref. | Ref. | Ref. | 77 / 164 (47%) | Ref. | Ref. |
| Any FIASMA<br>psychotropic<br>medication | 57 / 164 (34.8%) | 0.42 (0.31 - 0.57;<br><0.001*) | 0.49 (0.36 - 0.67;<br><0.001*) | 0.50 (0.37 - 0.67;<br><0.001*) | 57 / 164 (34.8%) | 0.65 (0.45 - 0.93;<br>0.019*) | 0.55 (0.39 - 0.77;<br>0.001*) |

<sup>1</sup> Adjusted for age, sex, hospital, obesity, and number of medical conditions.
<sup>2</sup> Adjusted for sex and obesity.
\* Two-sided p-value is significant (p<0.05).
Abbreviations: HR, hazard ratio, CI confidence interval.

**Figure 2.**
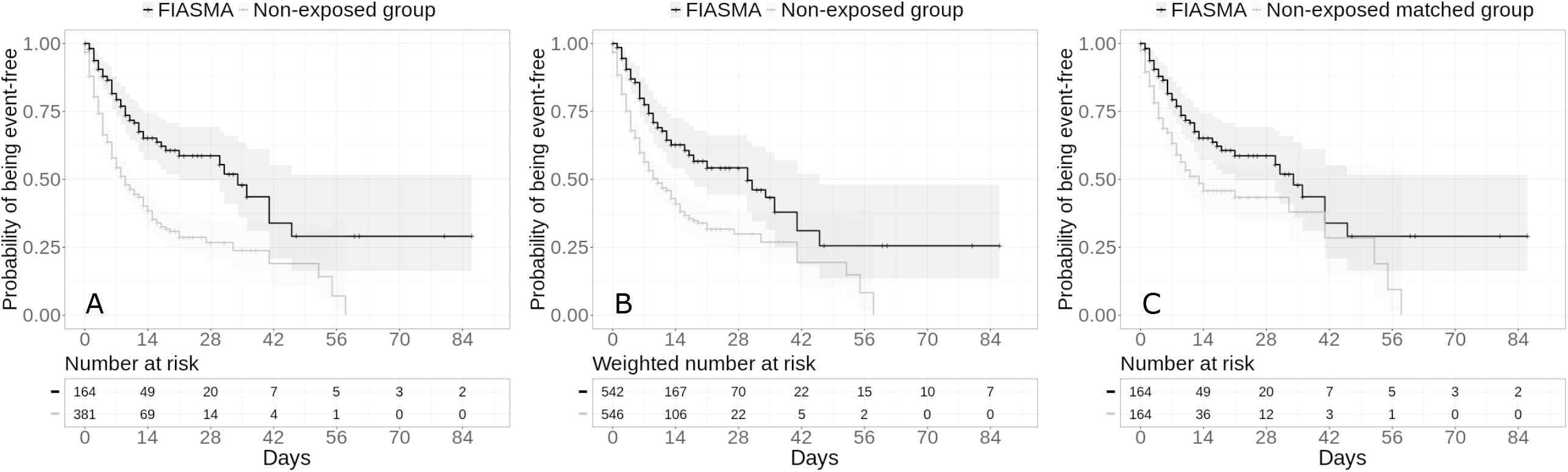
Kaplan-Meier curves for the composite endpoint of intubation or death in the full sample crude analysis (N=545) (A), in the full sample analysis with IPW (N=545) (B), and in the matched analytic sample using a 1:1 ratio (N=328) (C) among patients with mental disorder hospitalized for severe COVID-19, according to FIASMA psychotropic medication use at baseline.

In sensitivity analyses, the multivariable Cox regression model also yielded a significant association (HR=0.49; 95% CI=0.36-0.67; p<0.001), as did the Cox regression model in a matched analytic sample using a 1:1 ratio adjusted for unbalanced covariates, i.e. sex, and number of medical conditions (HR=0.55; 95% CI=0.39-0.77; p=0.001) (**Table 2**).

Additional exploratory analyses showed that FIASMA antidepressant use was significantly associated with reduced risk of intubation or death across all analyses (**Supplementary Table 3**). FIASMA antipsychotic use was significantly associated with reduced risk only in the multivariable Cox regression model and in the Cox regression model in a matched analytic sample using a 1:2 ratio adjusted for unbalanced covariates, possibly because of limited statistical power due to the limited number of patients receiving a FIASMA antipsychotic at hospital admission (N=13) (**Supplementary Table 3**). Hazard ratios were lower than 1 for most individual FIASMA molecules, but none of them reached statistical significance across all main and sensitivity analyses, except for hydroxyzine and escitalopram, possibly because of limited statistical power due to individual sample sizes ≤42 patients. Patients receiving a FIASMA antidepressant at baseline (N=148) had a significantly reduced risk of intubation or death compared with those receiving a non-FIASMA antidepressant at baseline (N=83) (**Supplementary Table 4; Supplementary Figure 1**). Adjusted analyses between FIASMA and non-FIASMA antipsychotics and antihistaminic medications could not be performed due to the insufficient number of events (i.e., <5) in the FIASMA groups (**Supplementary Table 4**). Finally, reproducing the main analyses among all patients with mental disorder with and without clinical severity criteria at baseline did not alter the significance of our results (**Supplementary Table 5**), as for the main analyses considering venlafaxine, mirtazapine and citalopram as FIASMA antidepressants (**Supplementary Table 6**).

## 4. Discussion

In this multicenter retrospective observational study involving a relatively large sample of patients with mental disorder hospitalized for severe COVID-19 (N=545), we found that FIASMA psychotropic medication use at study baseline was significantly and substantially associated with reduced risk of intubation or death, independently of sociodemographic characteristics, psychiatric and other medical comorbidity, and psychotropic and other medications. This association remained significant in multiple sensitivity analyses. Secondary exploratory analyses suggested that this association was not specific to one FIASMA psychotropic class or medication in this population. These results suggest that the acid sphingomyelinase (ASM)/ceramide system may provide a useful framework for the repurposing of FIASMA psychotropic medications against COVID-19 among individuals with mental disorders. Our findings also support the need of double-blind controlled randomized clinical trials of these medications for COVID-19 in this population, and more broadly in patients with severe COVID-19.

We found that FIASMA psychotropic medication use was significantly and substantially associated with reduced risk of intubation or death among patients with mental disorder hospitalized for severe COVID-19, and that this association was not specific to one FIASMA psychotropic class or medication in this population. These findings are in line with prior preclinical [4, 6] and clinical [7–9] evidence that FIASMA antidepressant medications may substantially prevent cells from being infected with SARS-CoV-2 *in vitro* [4, 6], and that FIASMA antidepressant medications and the FIASMA hydroxyzine at their usual respective antidepressant and antihistaminic doses, may reduce mortality among patients hospitalized for COVID-19 [7–9].

However, several alternative mechanisms could be proposed to explain this association. First, antiviral effects, i.e. inhibition of viral replication, of FIASMA medications might underlie this relationship, as suggested by a recent in-vitro study [4] for fluoxetine. However, inhibition of viral replication was not observed for other selective serotonin reuptake inhibitors (SSRIs) and FIASMA medications, including paroxetine and escitalopram. Second, many antidepressants (except for example sertraline and paroxetine) have high affinity for Sigma-1 receptors (S1R) [30, 31], and SSRIs have been suggested to have potential value in regulating inflammation by inhibiting cytokine production in COVID-19 [7, 32]. Because most FIASMA psychotropic antidepressants are S1R agonists, this mechanism might have overlapped their inhibition effect on ASM. However, when examining the association between several FIASMA psychotropic medications with low or no affinity for S1R (i.e., sertraline, paroxetine, duloxetine, aripiprazole and chlorpromazine) [30, 33–35] and the endpoint, main results remained statistically significant (**Supplementary Table 7**), suggesting that inhibition of ASM could underlie this association independently of S1R. Finally, this association may be partly mediated by the anti-inflammatory effects of FIASMA psychotropic medications, which could be explained by inhibition of the ASM in endothelial cells and the immune system, and might be independent of Sigma-1 receptors. First, a recent meta-analysis [36] of studies conducted in individuals with major depressive disorder following antidepressant treatment, mostly including SSRIs, supports that, overall, antidepressants may be associated with decreased plasma levels of 4 of 16 tested inflammatory mediators, including IL-10, TNF-α, and CCL-2, which are associated with COVID-19 severity [37], as well as IL-6, which is highly correlated with disease mortality [37, 38]. Second, prior *in vitro* and *in vivo* studies [39–41] suggest that antipsychotics may induce anti-inflammatory effect dependently on glia activation, and that this activity may not be shared by all antipsychotics. However, this anti-inflammatory effect was observed for both FIASMA antipsychotics (e.g. chlorpromazine) and non-FIASMA ones (e.g., haloperidol and risperidone). If the association between FIASMA psychotropic medication use and reduced risk of intubation or death were confirmed, future studies aiming at disentangling these potentially interrelated mechanisms are needed.

Our study has several limitations. First, there are two possible major inherent biases in observational studies: unmeasured confounding and confounding by indication. We tried to minimize the effects of confounding in several different ways. First, we used an analysis with inverse probability weighting to minimize the effects of confounding by indication [23, 24]. Second, we performed multiple sensitivity analyses, which showed similar results. Finally, although some amount of unmeasured confounding may remain, our analyses adjusted for numerous potential confounders. Other limitations include missing data for some baseline characteristic variables (i.e., 11.5%), which might be explained by the overwhelming of all hospital units during the COVID-19 peak incidence, and different results might have been observed during a lower COVID-19 incidence period. However, imputation of missing data did not alter the significance of our results (data available on request). Second, inflation of type I error might have occurred in secondary exploratory analyses due to multiple testing. Third, data on several FIASMA psychotropic medications, such as fluvoxamine or maprotiline, were not available because no patients with mental disorder hospitalized for severe COVID-19 received them at study baseline in AP-HP hospitals. Fourth, this study cannot establish a causal relationship between FIASMA psychotropic medication use and reduced risk of intubation or death [42]. Finally, despite the multicenter design, our results may not be generalizable to outpatients or other regions.

In this multicenter observational retrospective study, FIASMA psychotropic medication use at baseline was significantly associated with a reduced risk of intubation or death among adult patients with mental disorder hospitalized for severe COVID-19. These findings suggest the usefulness of the ASM/ceramide system framework in COVID-19. Double-blind controlled randomized clinical trials of these medications for COVID-19 are needed, starting with FIASMA molecules with the highest *in vitro* inhibition effect on ASM and the highest effect size in our study, and ease of use, including high safety margin, good tolerability, widespread availability, and low cost such that primary care physicians and other providers could prescribe them if their usefulness in patients with COVID-19 were confirmed.

## Data Availability

Data from the AP-HP Health Data Warehouse can be obtained with permission at https://eds.aphp.fr//.

## ACKNOWLEDGMENT SECTION

### Contributions

NH designed the study, performed statistical analyses, and wrote the first draft of the manuscript. MSR contributed to study design, performed statistical analyses and critically revised the manuscript. FL contributed to study design and critically revised the manuscript for scientific content. MA and PM performed statistical analyses and critically revised the manuscript. RV contributed to statistical analyses and critically revised the manuscript for scientific content. All other authors critically revised the manuscript for scientific content.

## Data Availability Statement

Data from the AP-HP Health Data Warehouse can be obtained with permission at https://eds.aphp.fr//.

## Disclaimer

The information contained in this study is provided for research purpose and should not be used as a substitute or replacement for diagnosis or treatment recommendations or other clinical decisions or judgment.

## Acknowledgments

The authors thank the EDS APHP Covid consortium integrating the APHP Health Data Warehouse team as well as all the APHP staff and volunteers who contributed to the implementation of the EDS-Covid database and operating solutions for this database. Collaborators of the EDS APHP Covid consortium: Pierre-Yves ANCEL, Alain BAUCHET, Nathanaël BEEKER, Vincent BENOIT, Mélodie BERNAUX, Ali BELLAMINE, Romain BEY, Aurélie BOURMAUD, Stéphane BREANT, Anita BURGUN, Fabrice CARRAT, Charlotte CAUCHETEUX, Julien CHAMP, Sylvie CORMONT, Christel DANIEL, Julien DUBIEL, Catherine DUCLOAS, Loic ESTEVE, Marie FRANK, Nicolas GARCELON, Alexandre GRAMFORT, Nicolas GRIFFON, Olivier GRISEL, Martin GUILBAUD, Claire HASSEN-KHODJA, François HEMERY, Martin HILKA, Anne Sophie JANNOT, Jerome LAMBERT, Richard LAYESE, Judith LEBLANC, Léo LEBOUTER, Guillaume LEMAITRE, Damien LEPROVOST, Ivan LERNER, Kankoe LEVI SALLAH, Aurélien MAIRE, Marie-France MAMZER, Patricia MARTEL, Arthur MENSCH, Thomas MOREAU, Antoine NEURAZ, Nina ORLOVA, Nicolas PARIS, Bastien RANCE, Hélène RAVERA, Antoine ROZES, Elisa SALAMANCA, Arnaud SANDRIN, Patricia SERRE, Xavier TANNIER, Jean-Marc TRELUYER, Damien VAN GYSEL, Gaël VAROQUAUX, Jill Jen VIE, Maxime WACK, Perceval WAJSBURT, Demian WASSERMANN, Eric ZAPLETAL.

