## Supplementary material for "Association between Psychotropic Medications Functionally Inhibiting Acid Sphingomyelinase and reduced risk of Intubation or Death among Individuals with Mental Disorder and Severe COVID-19: an Observational Study"

### Supplementary Text

### Variable assessment

We obtained the following data for each patient at the time of the hospitalization: sex; age, which was categorized into 4 classes based on the OpenSAFELY study results[20] (i.e. 18-50, 51-70, 71-80, 81+); hospital, which was categorized into 4 classes following the administrative clustering of AP-HP hospitals in Paris and its suburbs based on their geographical location (i.e., AP-HP Centre – Paris University, Henri Mondor University Hospitals and at home hospitalization; AP-HP Nord and Hôpitaux Universitaires Paris Seine-Saint-Denis; AP-HP Paris Saclay University; and AP-HP Sorbonne University); obesity, which was defined as having a body mass index higher than 30 kg/m2 or an ICD-10 diagnosis code for obesity (E66.0, E66.1, E66.2, E66.8, E66.9); self-reported current smoking status; number of medical conditions associated with severe COVID-19 [20–23], based on ICD-10 diagnosis codes, including diabetes mellitus (E11), diseases of the circulatory system (I00-I99), diseases of the respiratory system (J00-J99), neoplasms (C00-D49), and diseases of the blood and blood-forming organs and certain disorders involving the immune mechanism (D5-D8); and any medication prescribed according to compassionate use or as part of a clinical trial (e.g. hydroxychloroquine, azithromycin, remdesivir, tocilizumab, sarilumab, or dexamethasone). To take into account possible confounding by indication bias for psychotropic medications, we recorded whether patients had any current diagnosis of substance use disorder (F10-F19), psychotic disorder (F20-F29), mood or anxiety disorder (F30-F48), delirium or dementia (F05, R41.0 and G31.0), or any other psychiatric disorder (F01-F04, F06-F09 and F50-F99) based on ICD-10 diagnosis codes, and whether they were prescribed any antidepressant, mood stabilizer, benzodiazepine or Z-drug, or antipsychotic medication.

All medical notes and prescriptions are computerized in Greater Paris University hospitals. Medications including their dosage, frequency, date, and mode of administration were identified from medication administration data or scanned hand-written medical prescriptions, through two deep learning models based on BERT contextual embeddings [43], one for the medications and another for their mode of administration. The model was trained on the APmed corpus [44], a previously annotated dataset for this task. Extracted medications names were then normalized to the Anatomical Therapeutic Chemical (ATC) terminology using approximate string matching.

**Supplementary Figure 1. Kaplan-Meier curves for the composite endpoint of intubation or death in the full sample crude analysis (N=231) (A), in the full sample analysis with IPW (N=231) (B), and in the matched analytic sample using a 1:1 ratio (N=166) (C) among patients with mental disorder hospitalized for severe COVID-19 and receiving a FIASMA *versus* a non-FIASMA antidepressant at baseline.**

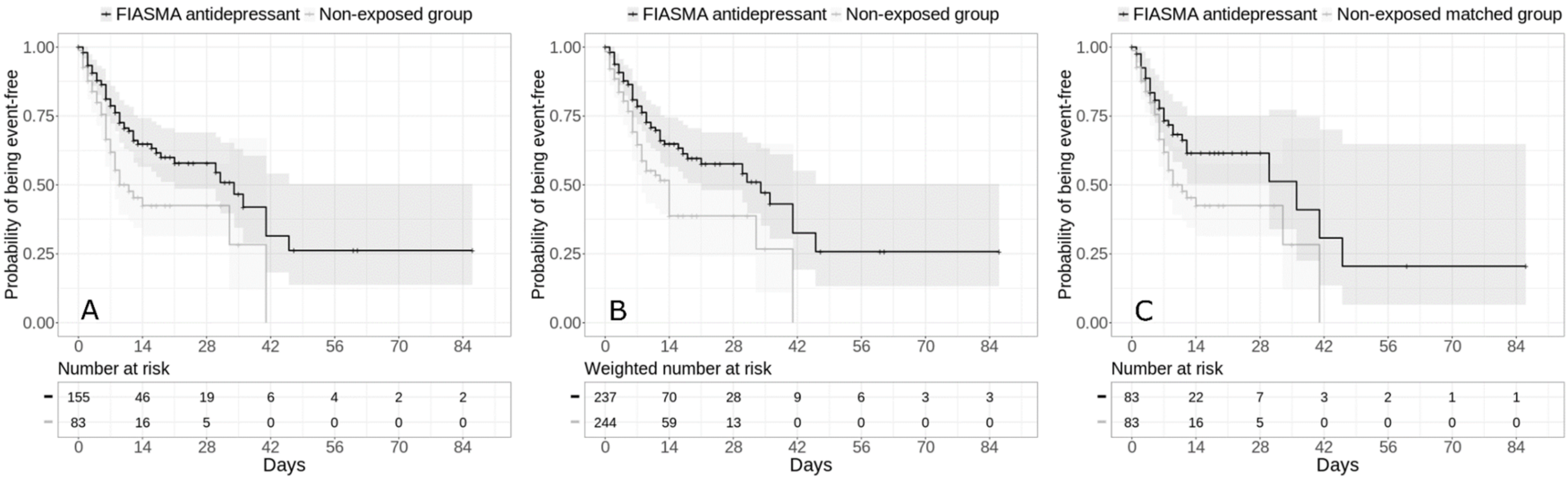


The shaded areas represent pointwise 95% confidence intervals.

**Supplementary Table 1. Associations of baseline clinical characteristics with the composite endpoint of intubation or death in the cohort of adult patients with mental disorder who had been admitted to AP-HP hospitals for severe COVID-19.**

| | Full sample (N=545) | With the endpoint (N=272) | Without the endpoint (N=273) | Crude analysis |
|---|---|---|---|---|
| | N (%) | N (%) | N (%) | HR (95% CI; p-value) |
| Age | | | | |
| *18 to 50 years* | 45 (8.26%) | 15 (33.3%) | 30 (66.7%) | Ref. |
| *51 to 70 years* | 148 (27.2%) | 57 (38.5%) | 91 (61.5%) | 0.98 (0.56 - 1.74; 0.956) |
| *71 to 80 years* | 112 (20.6%) | 60 (53.6%) | 52 (46.4%) | 1.53 (0.87 - 2.69; 0.143) |
| *More than 80 years* | 240 (44.0%) | 140 (58.3%) | 100 (41.7%) | 1.64 (0.96 - 2.79; 0.069) |
| Sex | | | | |
| *Women* | 250 (45.9%) | 110 (44.0%) | 140 (56.0%) | Ref. |
| *Men* | 295 (54.1%) | 162 (54.9%) | 133 (45.1%) | 1.42 (1.11 - 1.81; 0.005*) |
| Hospital | | | | |
| *AP-HP Centre – Paris University, Henri Mondor University Hospitals and at home hospitalization* | 151 (27.7%) | 90 (59.6%) | 61 (40.4%) | Ref. |
| *AP-HP Nord and Hôpitaux Universitaires Paris Seine-Saint-Denis* | 122 (22.4%) | 50 (41.0%) | 72 (59.0%) | 0.65 (0.46 - 0.92; 0.016*) |
| *AP-HP Paris Saclay University* | 154 (28.3%) | 78 (50.6%) | 76 (49.4%) | 0.73 (0.54 - 0.98; 0.039*) |
| *AP-HP Sorbonne University* | 118 (21.7%) | 54 (45.8%) | 64 (54.2%) | 0.65 (0.46 - 0.91; 0.013*) |
| Obesity [a] | | | | |
| *Yes* | 119 (21.8%) | 60 (50.4%) | 59 (49.6%) | 1.00 (0.75 - 1.34; 0.987) |

| | | | | |
|---|---|---|---|---|
| *No* | 426 (78.2%) | 212 (49.8%) | 214 (50.2%) | Ref. |
| Smoking [b] | | | | |
| *Yes* | 118 (21.7%) | 62 (52.5%) | 56 (47.5%) | 0.87 (0.65 - 1.16; 0.350) |
| *No* | 427 (78.3%) | 210 (49.2%) | 217 (50.8%) | Ref. |
| Number of medical conditions [c] | | | | |
| *0* | 93 (17.1%) | 18 (19.4%) | 75 (80.6%) | Ref. |
| *1* | 54 (9.91%) | 18 (33.3%) | 36 (66.7%) | 2.17 (1.13 - 4.17; 0.021*) |
| *2 or more* | 398 (73.0%) | 236 (59.3%) | 162 (40.7%) | 4.00 (2.47 - 6.46; <0.001*) |
| Any current substance abuse disorder [d] | | | | |
| *Yes* | 67 (12.3%) | 38 (56.7%) | 29 (43.3%) | 1.37 (0.97 - 1.92; 0.075) |
| *No* | 478 (87.7%) | 234 (49.0%) | 244 (51.0%) | Ref. |
| Any current psychotic disorder [e] | | | | |
| *Yes* | 31 (5.69%) | 13 (41.9%) | 18 (58.1%) | 0.78 (0.44 - 1.35; 0.371) |
| *No* | 514 (94.3%) | 259 (50.4%) | 255 (49.6%) | Ref. |
| Any current mood or anxiety disorder [f] | | | | |
| *Yes* | 93 (17.1%) | 52 (55.9%) | 41 (44.1%) | 1.08 (0.80 - 1.46; 0.632) |
| *No* | 452 (82.9%) | 220 (48.7%) | 232 (51.3%) | Ref. |
| Delirium or dementia [g] | | | | |
| *Yes* | 153 (28.1%) | 99 (64.7%) | 54 (35.3%) | 1.51 (1.18 - 1.94; 0.001*) |
| *No* | 392 (71.9%) | 173 (44.1%) | 219 (55.9%) | Ref. |
| Any other mental disorder [h] | | | | |
| *Yes* | 43 (7.89%) | 28 (65.1%) | 15 (34.9%) | 1.79 (1.21 - 2.65; 0.004*) |
| *No* | 502 (92.1%) | 244 (48.6%) | 258 (51.4%) | Ref. |
| Medication according to compassionate use or as part of a clinical trial [i] | | | | |
| *Yes* | 142 (26.1%) | 66 (46.5%) | 76 (53.5%) | 0.88 (0.66 - 1.16; 0.349) |
| *No* | 403 (73.9%) | 206 (51.1%) | 197 (48.9%) | Ref. |

| | | | | |
|---|---|---|---|---|
| Any antidepressant | | | | |
| *Yes* | 238 (43.7%) | 95 (39.9%) | 143 (60.1%) | 0.53 (0.42 - 0.68; <0.001*) |
| *No* | 307 (56.3%) | 177 (57.7%) | 130 (42.3%) | Ref. |
| Any antipsychotic medication | | | | |
| *Yes* | 93 (17.1%) | 30 (32.3%) | 63 (67.7%) | 0.56 (0.38 - 0.82; 0.003*) |
| *No* | 452 (82.9%) | 242 (53.5%) | 210 (46.5%) | Ref. |
| Any mood stabilizer medication [j] | | | | |
| *Yes* | 92 (16.9%) | 34 (37.0%) | 58 (63.0%) | 0.64 (0.45 - 0.92; 0.016*) |
| *No* | 453 (83.1%) | 238 (52.5%) | 215 (47.5%) | Ref. |
| Any benzodiazepine or Z-drug | | | | |
| *Yes* | 239 (43.9%) | 122 (51.0%) | 117 (49.0%) | 0.87 (0.68 - 1.11; 0.251) |
| *No* | 306 (56.1%) | 150 (49.0%) | 156 (51.0%) | Ref. |

[a] Defined as having a body-mass index higher than 30 kg/m$^2$ or an International Statistical Classification of Diseases and Related Health Problems (ICD-10) diagnosis code for obesity (E66.0, E66.1, E66.2, E66.8, E66.9).
[b] Current smoking status was self-reported.
[c] Assessed using ICD-10 diagnosis codes for diabetes mellitus (E11), diseases of the circulatory system (I00-I99), diseases of the respiratory system (J00-J99), neoplasms (C00-D49), diseases of the blood and blood-forming organs and certain disorders involving the immune mechanism (D5-D8), frontotemporal dementia (G31.0), peptic ulcer (K27), diseases of liver (K70-K95), hemiplegia or paraplegia (G81-G82), acute kidney failure or chronic kidney disease (N17-N19), and HIV (B20).
[d] Assessed using ICD-10 diagnosis codes for mental and behavioural disorders due to psychoactive substance use (F10-F19).
[e] Assessed using ICD-10 diagnosis codes for psychotic disorders (F20-F29).
[f] Assessed using ICD-10 diagnosis codes for mood and anxiety disorders (F30-F48).
[g] Assessed using ICD-10 diagnosis codes for delirium or dementia (F05, R41.0 and G31.0).
[h] Assessed using ICD-10 diagnosis codes for other psychiatric disorders (F01-F04, F06-F09 and F50-F99).
[i] Any medication prescribed as part of a clinical trial or according to compassionate use (e.g., hydroxychloroquine, azithromycin, remdesivir, tocilizumab, sarilumab, or dexamethasone).
[j] Included lithium or antiepileptic medications with mood stabilizing properties.
* Two-sided p-value is significant (p<0.05).
Abbreviations: HR, hazard ratio, CI confidence interval.

**Supplementary Table 2. Backward stepwise regression models.**

| Step 1 | AIC |
|---|---|
| Delirium or dementia, Any antidepressant , Any benzodiazepine or Z-drug , Any other mental disorder, Any current substance abuse disorder, Medication according to compassionate use or as part of a clinical trial, Any current psychotic disorder, Any mood stabilizer medication, Any antipsychotic, Smoking, Any current mood or anxiety disorder, Hospital, Obesity, Sex, Any FIASMA, Age, Number of conditions | 2976.87 |
| **Step 2** | |
| Delirium or dementia, Any benzodiazepine or Z-drug, Any other mental disorder, Any current substance abuse disorder, Medication according to compassionate use or as part of a clinical trial, Any current psychotic disorder, Any mood stabilizer medication, Any antipsychotic, Smoking, Any current mood or anxiety disorder, Hospital, Obesity, Sex, Any FIASMA, Age, Number of conditions | 2974.87 |
| **Step 3** | |
| Any benzodiazepine or Z-drug, Any other mental disorder, Any current substance abuse disorder, Medication according to compassionate use or as part of a clinical trial, Any current psychotic disorder, Any mood stabilizer medication, Any antipsychotic, Smoking, Any current mood or anxiety disorder, Hospital, Obesity, Sex, Age, Any FIASMA, Number of conditions | 2972.88 |
| **Step 4** | |
| Any other mental disorder, Any current substance abuse disorder, Medication according to compassionate use or as part of a clinical trial, Any current psychotic disorder, Any antipsychotic, Any mood stabilizer medication, Smoking, Any current mood or anxiety disorder, Hospital, Obesity, Sex, Age, Any FIASMA, Number of conditions | 2970.91 |
| **Step 5** | |
| Any current substance abuse disorder, Medication according to compassionate use or as part of a clinical trial, Any antipsychotic, Any current psychotic disorder, Any mood stabilizer medication, Smoking, Any current mood or anxiety disorder, Hospital, Obesity, Sex, Age, Any FIASMA, Number of conditions | 2969 |
| **Step 6** | |
| Medication according to compassionate use or as part of a clinical trial, Any antipsychotic, Any current psychotic disorder, Smoking, Any mood stabilizer medication, Any current mood or anxiety disorder, Hospital, Obesity, Sex, Age, Any FIASMA, Number of conditions | 2967.12 |
| **Step 7** | |
| Any antipsychotic, Any current psychotic disorder, Any mood stabilizer medication, Smoking, Any current mood or anxiety disorder, Hospital, Obesity, Sex, Age, Any FIASMA, Number of conditions | 2965.54 |
| **Step 8** | |
| Smoking, Any mood stabilizer medication, Any antipsychotic, Any current mood or anxiety disorder, Hospital, Obesity, Sex, Age, Any FIASMA, Number of conditions | 2964.25 |

| | |
|---|---|
| **Step 9** | |
| Smoking, Any antipsychotic, Any current mood or anxiety disorder, Hospital, Obesity, Sex, Age, Any FIASMA, Number of conditions | 2962.95 |
| **Step 10** | |
| Any antipsychotic, Any current mood or anxiety disorder, Hospital, Obesity, Sex, Age, Any FIASMA, Number of conditions | 2961.68 |
| **Step 11** | |
| Any current mood or anxiety disorder, Hospital, Obesity, Sex, Age, Any FIASMA, Number of conditions | 2960.77 |
| **Step 12** | |
| Hospital, Obesity, Sex, Age, Any FIASMA, Number of conditions | 2959.87 |

**Supplementary Table 3. Association of individual FIASMA psychotropic classes and molecules at baseline with the risk of intubation or death among patients with mental disorder hospitalized for severe COVID-19.**

| | Number of events / Number of patients | Crude Cox regression analysis | Multivariable Cox regression analysis [a] | Analysis weighted by inverse-probability-weighting weights [a] | Analysis weighted by inverse-probability-weighting weights [a] adjusted for unbalanced covariates | Number of events / Number of patients in the matched control groups | Univariate Cox regression in a 1:2 ratio matched analytic sample [a] | Cox regression in a 1:2 ratio matched analytic sample[a] adjusted for unbalanced covariates |
|---|---|---|---|---|---|---|---|---|
| | N (%) | HR (95% CI; p-value) | HR (95% CI; p-value) | HR (95% CI; p-value) | HR (95% CI; p-value) | N (%) | HR (95% CI; p-value) | HR (95% CI; p-value) |
| *No FIASMA psychotropic medication* | 215 / 381 (56.4) | Ref. | Ref. | Ref. | Ref. | Ref. | Ref. | Ref. |
| *FIASMA Antidepressants* | 53 / 148 (35.8) | 0.44 (0.33 – 0.60; <0.001*) | 0.52 (0.8 – 0.71; <0.001*) | 0.51 (0.38 – 0.69; <0.001*) | - | 160 / 296 (54.1) | 0.47 (0.34 – 0.66; <0.001*) | 0.50 (0.37 – 0.69; <0.001*) [b] |
| Amitriptyline | 8 / 20 (40.0) | 0.43 (0.21 – 0.88; 0.021*) | 0.44 (0.20 – 0.95 0.036*) | 0.76 (0.38 – 1.50; 0.426) | 0.70 (0.37 – 1.30; 0.252) [c] | 21 / 40 (52.5) | 0.52 (0.23 – 1.19; 0.123) | 0.55 (0.23 – 1.32; 0.183) [d] |
| Clomipramine | 1 / 4 (25.0) | 0.46 (0.07 – 3.30; 0.441) | NA | NA | NA | NA | NA | NA |
| Duloxetine | 4 / 11 (36.4) | 0.38 (0.14 – 1.03; 0.056) | NA | NA | NA | NA | NA | NA |
| Escitalopram | 12 / 42 (28.6) | 0.35 (0.20 – 0.63; <0.001*) | 0.61 (0.37 – 1.01 ; 0.005*) | 0.39 (0.21 – 0.70 ; 0.002*) | 0.34 (0.18 – 0.63; <0.001*) [e] | 41 / 84 (48.8) | 0.42 (0.22 – 0.80; 0.009*) | 0.40 (0.21 – 0.76 ; 0.006*) [f] |
| Fluoxetine | 4 / 14 (28.6) | 0.29 (0.11 – 0.79 ; 0.015*) | NA | NA | NA | NA | NA | NA |
| Paroxetine | 19 / 44 (43.2) | 0.54 (0.33 – 0.86 ; 0.010*) | 0.65 (0.39 – 1.04 ; 0.007*) | 0.85 (0.54 – 1.35; 0.495) | 0.77 (0.48 – 1.23; 0.272) [g] | 43 / 88 (48.9) | 0.63 (0.36 – 1.09 ; 0.097) | 0.61 (0.35 – 1.06 ; 0.078) [h] |
| Sertraline | 7 / 21 (33.3) | 0.41 (0.19 – 0.86 ; 0.019*) | 0.47 (0.24 – 0.93 ; 0.029*) | 0.57 (0.28 – 1.18 ; 0.129) | 0.50 (0.21 – 1.20 ; 0.119) [i] | 15 / 42 (35.7) | 0.72 (0.29 – 1.76 ; 0.468) | 0.77 (0.31 – 1.89 ; 0.564) [j] |
| *FIASMA Antipsychotics* | 4 / 13 (30.8) | 0.38 (0.14 – 1.03; 0.058) | 0.40 (0.17 – 0.97; 0.042*) | 0.58 (0.23 – 1.47; 0.249) | 0.58 (0.19 – 1.72; 0.324) [k] | 12 / 26 (46.2) | 0.42 (0.13 – 1.32; 0.139) | 0.25 (0.07 - 0.90; 0.034*) |
| Aripiprazole | 1 / 7 (14.3) | 0.16 (0.02 – 1.17; 0.071) | NA | NA | NA | NA | NA | NA |

| | | | | | | | | |
|---|---|---|---|---|---|---|---|---|
| Chlorpromazine | 3 / 6 (50.0) | 0.75 (0.35 – 1.61; 0.457) | NA | NA | NA | NA | NA | NA |
| *FIASMA Antihistaminics* | | | | | | | | |
| Hydroxyzine | 4 / 17 (23.5) | 0.21 (0.08 – 0.58; 0.002*) | 0.16 (0.05 – 0.55; 0.004*) | 0.17 (0.03 – 0.84; 0.030*) | 0.26 (0.09 – 0.72; 0.010*) [l] | 18 / 34 (52.9) | 0.27 (0.09 – 0.82; 0.020*) | 0.17 (0.05 – 0.57; 0.005*) [m] |

[a] Adjusted for age, sex, hospital, obesity, and number of medical conditions.
[b] Adjusted for sex, hospital, and number of medical conditions.
[c] Adjusted for age, hospital, and number of medical conditions.
[d] Adjusted for sex and hospital.
[e] Adjusted for age and number of medical conditions.
[f] Adjusted for sex and hospital.
[g] Adjusted for sex, hospital, and number of medical conditions.
[h] Adjusted for sex and obesity.
[i] Adjusted for sex and number of medical conditions.
[j] Adjusted for sex and obesity.
[k] Adjusted for age, sex, hospital, obesity, and number of medical conditions.
[l] Adjusted for age, sex, hospital, obesity, and number of medical conditions.
[m] Adjusted for sex and hospital.
* Two-sided p-value is significant (p<0.05).
Abbreviations: HR, hazard ratio, CI confidence interval, NA, not applicable.

**Supplementary Table 4. Association between FIASMA and non-FIASMA psychotropic medication use and the endpoint of intubation or death among patients with mental disorder hospitalized for severe COVID-19.**

| | **Number of events / Number of patients** | **Crude Cox regression analysis** | **Multivariable Cox regression analysis** [a] | **Analysis weighted by inverse-probability-weighting weights** [a] | **Analysis weighted by inverse-probability-weighting weights** [a] **adjusted for unbalanced covariates** | **Number of events / Number of patients in the matched groups** | **Univariate Cox regression in a 1:2 ratio matched analytic sample** [a] | **Cox regression in a 1:2 ratio matched analytic sample**[a] **adjusted for unbalanced covariates** |
|---|---|---|---|---|---|---|---|---|
| | N (%) | HR (95%CI; p-value) | HR (95%CI; p-value) | HR (95%CI; p-value) | HR (95%CI; p-value) | N (%) | HR (95%CI; p-value) | HR (95%CI; p-value) |
| FIASMA antidepressants | 53 / 148 (35.8) | 0.53 (0.35 – 0.81 ; 0.003*) | 0.45 (0.29 – 0.71 ; <0.001*) | 0.55 (0.36 – 0.83; 0.005*) | - | 31 / 83 (37.3) | 0.61 (0.38 – 0.98; 0.042*) | 0.55 (0.34 – 0.89; 0.016*) [b] |
| Other antidepressants | 40 / 83 (48.2) | Ref. | Ref. | Ref. | Ref. | 340/ 83 (480.2) | Ref. | Ref. |
| FIASMA antipsychotics | 4 / 13 (30.8) | 0.97 (0.367 – 2.53; 0.947) | NA | NA | NA | 4 / 13 (30.8) | 0.57 (0.17 – 1.89; 0.356) | NA |
| Other antipsychotics | 12 / 45 (26.7) | Ref. | Ref. | Ref. | NA | 5 / 13 (38.5) | Ref. | NA |
| FIASMA antihistaminics | 4 / 17 (23.5) | 1.01 (0.11 – 9.13; 0.99) | NA | NA | NA | NA | NA | NA |
| Other antihistaminics | 1 / 5 (20.0) | Ref. | Ref. | Ref. | Ref. | NA | Ref. | Ref. |

[a] Adjusted for age, sex, hospital, obesity, and number of medical conditions.
[b] Adjusted for number of medical conditions.
* Two-sided p-value is significant (p<0.05).
Abbreviations: HR, hazard ratio, CI confidence interval, NA, not applicable.

**Supplementary Table 5. Association between FIASMA psychotropic medication use at baseline and intubation or death in patients with mental disorder hospitalized for COVID-19 with and without clinical severity of COVID-19 at admission (N=1,406).**

| | Number of events / Number of patients | Crude Cox regression analysis | Multivariable Cox regression analysis [a] | Analysis weighted by inverse-probability-weighting weights | Analysis weighted by inverse-probability-weighting weights [a] adjusted for unbalanced covariates | Number of events / Number of patients in the matched groups | Univariate Cox regression in a 1:1 ratio matched analytic sample [a] | Cox regression in a 1:1 ratio matched analytic sample [a] adjusted for unbalanced covariates |
|---|---|---|---|---|---|---|---|---|
| | N (%) | HR (95% CI; p-value) | HR (95% CI; p-value) | HR (95% CI; p-value) | HR (95% CI; p-value) | N (%) | HR (95% CI; p-value) | HR (95% CI; p-value) |
| No FIASMA psychotropic medication | 15 / 902 (38.9) | Ref. | Ref. | Ref. | Ref. | 161 / 504 (31.9) | Ref. | Ref. |
| Any FIASMA psychotropic medication | 351 / 504 (22.6) | 0.47 (0.38 – 0.58; <0.001*) | 0.67 (0.52 – 0.87; <0.001*) | 0.63 (0.51 – 0.78; <0.001*) | NA | 114 / 504 (22.6) | 0.60 (0.48 – 0.77; <0.001*) | 0.64 (0.51 – 0.82; <0.001*) [b] |

[a] Adjusted for age, sex, hospital, obesity and number of medical conditions.
[b] Additionally adjusted for sex and number of medical conditions.
* Two-sided p-value is significant (p<0.05).
Abbreviations: HR, hazard ratio, CI confidence interval.

**Supplementary Table 6. Association between FIASMA psychotropic medication use at baseline and risk of intubation or death among patients with mental disorder hospitalized for severe COVID-19 while considering venlafaxine, mirtazapine and citalopram as FIASMA antidepressants.**

| | Number of events / Number of patients | Crude Cox regression analysis | Multivariable Cox regression analysis [a] | Analysis weighted by inverse-probability-weighting weights | Analysis weighted by inverse-probability-weighting weights [a] adjusted for unbalanced covariates [b] | Number of events / Number of patients in the matched groups | Univariate Cox regression in a 1:2 ratio matched analytic sample [a] | Cox regression in a 1:2 ratio matched analytic sample [a] adjusted for unbalanced covariates [c] |
|---|---|---|---|---|---|---|---|---|
| | N (%) | HR (95% CI; p-value) | HR (95% CI; p-value) | HR (95% CI; p-value) | HR (95% CI; p-value) | N (%) | HR (95% CI; p-value) | HR (95% CI; p-value) |
| No FIASMA psychotropic medication | 200 / 354 (56.5) | Ref. | Ref. | Ref. | Ref. | 92 / 191 (48.2) | Ref. | Ref. |
| Any FIASMA psychotropic medication | 72 / 191 (37.7) | 0.48 (0.37 – 0.63; <0.001*) | 0.55 (0.42 – 0.73; <0.001*) | 0.61 (0.46 – 0.80; <0.001*) | 0.54 (0.41 – 0.71; <0.001*) | 72 / 191 (37.7) | 0.59 (0.43 – 0.80; 0.001*) | 0.58 (0.43 – 0.80; 0.001*) |

[a] Adjusted for age, sex, hospital, obesity and number of medical conditions.
[b] Additionally adjusted for sex, and number of medical conditions.
[c] Additionally adjusted for sex, and obesity.
* Two-sided p-value is significant (p<0.05).
Abbreviations: HR, hazard ratio, CI confidence interval.

**Supplementary Table 7. Association between FIASMA psychotropic medications without Sigma-1-receptor agonist effect and intubation or death among patients with mental disorder hospitalized for severe COVID-19.**

| | Number of events / Number of patients | Crude Cox regression analysis | Multivariable Cox regression analysis [a] | Analysis weighted by inverse-probability-weighting weights | Analysis weighted by inverse-probability-weighting weights [a] adjusted for unbalanced covariates [b] | Number of events / Number of patients in the matched groups | Univariate Cox regression in a 1:2 ratio matched analytic sample [a] | Cox regression in a 1:2 ratio matched analytic sample [a] adjusted for unbalanced covariates [c] |
|---|---|---|---|---|---|---|---|---|
| | N (%) | HR (95% CI; p-value) | HR (95% CI; p-value) | HR (95% CI; p-value) | HR (95% CI; p-value) | N (%) | HR (95% CI; p-value) | HR (95% CI; p-value) |
| No FIASMA psychotropic medication | 215 / 381 (56.4) | Ref. | Ref. | Ref. | Ref. | 26 / 49 (53.1) | Ref. | Ref. |
| Any non S1R agonist and FIASMA psychotropic medication[d] | 15 / 49 (30.6) | 0.33 (0.20 – 0.57; <0.001*) | 0.30 (0.17 – 0.53; <0.001*) | 0.28 (0.27 – 0.76; 0.003*) | 0.37 (0.22 – 0.63; <0.001*) | 15 / 49 (30.6) | 0.39 (0.20 – 0.73; 0.004*) | 0.40 (0.21 – 0.77; 0.006*) |

[a] Adjusted for age, sex, hospital, obesity and number of medical conditions.
[b] Additionally adjusted for age, sex, hospital and number of medical conditions.
[c] Additionally adjusted for sex, and obesity.
[d] Any antipsychotic, hydroxyzine, Sertraline
* Two-sided p-value is significant (p<0.05).
Abbreviations: HR, hazard ratio, CI confidence interval.
